# Less is More: last observations of vital signs can outperform time series for hospital mortality prediction

**DOI:** 10.64898/2026.05.05.26352366

**Authors:** Zizheng Zhang, Jia Wei, Justin Xu, Yiming Li, Augustine Luk, Sejal Bhalla, Haochen Cui, David A Clifton, A Sarah Walker, David W Eyre

## Abstract

Timely identification of hospital inpatients at risk of deterioration facilitates interventions to support their recovery. Many hospitals implement early warning scores to detect abnormal patient vital signs, such as the National Early Warning Score 2 (NEWS2). However, these are typically based on a snapshot of the most recent vital signs, rather than exploiting trends overtime that clinical intuition suggests may also be informative. Multiple approaches, including recently described methods, have been developed to predict patient deterioration from time series. We therefore compared the effectiveness of different mortality prediction models, including clinical scoring systems, classical machine learning models and state-of-the-art deep learning models using both snapshot and time series vital sign data. No significant improvement in model performance was observed using predictions from time series compared to using the last observation of the time series and non-temporal features such as demographics. Our study comprehensively compares different model types, and provides recommendations for developing predictive models and guidance for what evaluation is needed before considering deploying such models in inpatient care.

## Introduction

Rapid and accurate early warning of patient deterioration can improve patient outcomes and reduce healthcare costs^1,2^. However, the high workload faced by healthcare staff, exacerbated by changes since the COVID-19 pandemic^3−6^, means capacity for manually monitoring patients may be limited^7^. In response, early warning systems, based on scoring how abnormal a patient’s vital signs are, have been developed. These use measurements of vital signs, such as heart rate, blood pressure and respiratory rate, to automate identification of patients at risk of adverse outcomes. In the UK, the National Early Warning Score 2 (NEWS2) has been widely deployed in hospitals for this purpose^8^, and achieves good performance^9,10^.

However, within such scoring systems there is a trade-off, where requiring high levels of sensitivity to support patient safety leads to relatively high ‘false alarm’ rates. Additionally, most scoring systems are based on a snapshot of the most recent vital signs, rather than making use of the trend over time that clinical intuition suggests may also be informative. Therefore, there has been considerable interest in trying to improve performance by applying machine learning techniques on vital signs data, and in particular time series of vital signs data, to predict patient adverse outcomes like mortality^11−16^.

Advances in artificial intelligence (AI) and deep learning have resulted in development of multiple new approaches for processing time series data. Vital signs of hospitalised patients form time series since patients’ vital signs are measured multiple times during an admission. Clinicians make use of the temporal patterns to assess a patient’s condition. However, most classical machine learning models are designed for tabular data and do not intrinsically integrate temporal dependencies across features. In contrast, several deep neural network (DNN) architectures can process sequential data potentially capturing trends leading to adverse events. These time series DNN architectures have achieved state-of-the-art performance for various tasks outside of healthcare on public benchmark datasets such as forecasting, classification, and interpolation^17−25^. Traditionally DNN time series approaches have used methods with a particular focus on models derived from recurrent neural networks, such as long short-term memory (LSTM) networks. However, a number of variants of Transformer model architectures have been adapted for time series processing by manipulating the attention mechanism^18^, or modifying the input structures of the time series, such as patching, which divides the time series to enable local pattern detection^19^. Convolution-based models like TimesNet^17^, which employs 2-dimensional temporal convolution to learn the seasonality of the time series, have also claimed state-of-the-art performance particularly in time series classification tasks. DLinear and FreTS^21,22^, which are based on Multi-Layer Perceptron networks, demonstrate competitive performance and offer computational efficiency benefiting from simpler architectures.

Despite these advances, there remains a lack of comprehensive evaluation of the comparative performance of DNN models, classical machine learning models, and early warning scores, at predicting mortality in a hospital-wide patient population. Rather, much previous research has focused on predicting mortality in intensive care (ICU)^25−27^, due to the availability of public datasets such as Medical Information Mart for Intensive Care (MIMIC)^12^; other specific settings have also been studied including Emergency Departments^16^. However, unselected medical and surgical admissions are the largest patient group in many hospitals and monitoring these patients requires significant human input from healthcare professionals. ICU patients are likely to be more unwell and on greater physiological support, compared to the general inpatient population, possibly leading to different underlying patterns. Furthermore, comparisons in the literature often focus on high-performing deep learning models and report excellent performance for these models, but may not consider existing early warning systems in use in hospitals^10^, or other established classical machine learning models, such as XGBoost, or clinician performance, as baselines^25,26^. Where comparisons have been made, several studies have identified that sophisticated deep learning with time series could not achieve higher performance than classical machine learning methods when predicting mortality in Emergency Departments, or in other nonmedical prediction tasks^28−30^. Even within deep learning models, looking over back longer periods of a time series does not always lead to better performance^31^. Additionally, previous literature has shown that when using tabular data, XGBoost outperforms most of the deep learning models across a range of tasks^29,32−34^. Together, this naturally leads to the question of whether more complex and sophisticated time series models lead to better performance when predicting patient outcomes in a broad range of hospitalised patients.

In this study, we therefore use large-scale inpatient data to undertake a comprehensive comparison of inpatient mortality prediction using established early warning scores, classical machine learning models, and multiple DNN architectures, in order to guide model selection and allow optimal utilisation of healthcare and computational resources. We also aim to promote greater standardisation of evaluations prior to deploying AI in hospital care.

## Methods

We used data from Oxford University Hospitals (OUH) NHS Foundation Trust, an organisation comprising 4 hospitals in Oxfordshire, UK, collectively containing ∼1100 beds serving ∼1% of the UK population as well as providing specialist regional referral services. Deidentified data covering patient demographics, mortality, hospital admissions, diagnostic codes and vital signs were obtained from the Infections in Oxfordshire Research Database, which has approvals from the National Research Ethics Service South Central-Oxford C Research Ethics Committee (24/SC/0241), Health Research Authority and Confidentiality Advisory Group (19/CAG/0144) as a deidentified database without individual consent.

Data were extracted for all adult inpatients (i.e. aged ≥ 16 years) who are admitted for planned or emergency care, between 01 January 2016 and 13 June 2024, excluding planned day cases, regular day/night admissions, and maternity admissions^35^. We used 7 vital signs and the date and time they were performed as model inputs (i.e. those also used in NEWS2): heart rate, respiratory rate, systolic blood pressure, oxygen saturation, level of consciousness (Table S1), temperature, and the use of supplemental oxygen. Vital sign measurements incompatible with life, i.e. reflecting manual data entry errors, were excluded from the dataset (Table S1). We also assessed the impact on model performance of including additional static model features including age, sex, ethnicity, co-morbidities (converting ICD10 diagnostic codes from admissions in the prior 1 year into domains of the Charlson comorbidity index), Index of Multiple Deprivation score (deprivation of each patient’s residential area), and admission method (emergency or elective, i.e. planned) (Table S2).

We aimed to predict mortality within the next 24 hours, using prior vital sign measurements as core input features for each model. Formally, this is a multivariable binary classification task, where a positive label indicates mortality within the next 24 hours. To generate datasets for training and testing models, we randomly selected 3,500 specific date-times (roughly one per day) between the start and end of the study period. At each time point, for each of the current hospital inpatients, we identified if the patient died within the next 24 hours. Given the different input requirements of each of the models evaluated, we applied different feature engineering as follows.

## Model architectures

### NEWS2

NEWS2 scores were calculated from the last available vital sign measurements. A logistic regression model was trained to predict 24-hour mortality using the NEWS2 score as the sole input feature. We also evaluated adding static non-vital sign features, such as age, in addition to the NEWS2 score, as inputs in separate models.

### XGBoost

*We* trained several models using an example classical machine learning model architecture, XGBoost. We evaluated several feature combinations: i) only vital sign values from the last available time point, ii) the full vital sign time series resampled hourly from the preceding 48 hours (the median was taken if there were multiple measurements within the hour) and iii) using the last recorded vital signs and the difference between the last and immediately preceding vital sign measurements. We also evaluated adding static information for each option described above. When modelling the full time series, hours without recorded measurements are treated as missing as XGBoost is inherently robust with missingness. We also evaluated linear interpolation between observed values, which did not result in a meaningful change in model performance (and so is not presented separately).

### DNN models

A selection of established and state-of-the-art deep learning model architectures, identified from existing benchmarks and surveys, were included to compare their performance on the dataset^32,36^. The underlying model architectures evaluated include: Convolutional Neural Network (CNN) models such as classical CNN, TimesNet^17^; Multi-layer Perceptron (MLP) models such as DLinear^21^; Recurrent Neural Networks (RNN) such as Long Short Term Memory networks (LSTM)^23^; and Transformer models such as vanilla Transformer, !Transformer and Crossformer^18,37,20^. To fit these DNN models, we used the preceding 48 hours of vital sign measurements as model inputs. Most require that the irregularly measured vital signs are transformed into a regularly sampled time series. We therefore resampled the vital signs every 60 minutes using linear interpolation. If the patient had less than 48 hours of history, i.e., was recently admitted, the missing values prior to this were imputed using the population median of the training set (without down sampling) of the corresponding vital signs (we also tested carrying the first observation backwards, which made minimal difference, see supplement for sensitivity analysis, Figure S2). The dataset was scaled using min-max scaling before proceeding to model training. Different length of lookback history, such as 24 hours or 12 hours, were also evaluated in a sensitivity analysis. Different interpolation strategies were also tested including last observation carried forwards (LOCF) and flexible model-based interpolation using Gaussian Processes; neither materially changed model performance. Previous benchmark studies using EHR datasets have shown that the model-based interpolation including deep learning-based interpolation generally yields a similar accuracy within the time series and on downstream tasks compared to linear interpolation and LOCF^32^. We also evaluated optionally incorporating non-temporal features such as age and sex, concatenating this static information with the output of sequential models in the DNN before passing it to an MLP-based classifier head.

The architecture of the models evaluated covers all typical deep learning base model types and was designed to learn the underlying patterns of the time series, including long-term and short-term dependencies. The implementation of the models either followed the official implementation released by the author of the original paper, or reimplementation from existing open source benchmark studies if the original model was not initially designed to be used for a classification task^32,36^.

### Model Training

The dataset was split randomly, stratified by patient pseudo-identifier, with 80% used for training, 10% used for validation (for selecting hyperparameters and model architectures) and 10% reserved as a held-out test set. Down-sampling was applied within the training data to balance the positive and negative outcomes given the significant class imbalance (many more patients survived than died in each time period). Validation and testing were performed without down-sampling to reflect real-world performance. Starting from recommended or default hyperpara meters^32,36^, random hyperparameter searching optimisations within plausible/recommended ranges were performed to maximise performance based on the original implementation of the model and prior benchmark studies^36^. While the number of models evaluated and size of the data prevented more exhaustive optimisation, 10 repeated search runs with multiple recommended initialisations for each model type failed to identify hyperparameters configurations yielding significant performance improvements before early stopping. All the data processing was performed using Python 3.11.5^38^, PyTorch 2.2.2^39^ was used for DNN model training. Scikit-learn 1.4.2^40^ and xgboost 2.1.2^41^ were used for logistic regression and XGBoost model training respectively. Further training details are included in the appendix.

### Performance evaluation

Model performance was evaluated in the held-out test data. Metrics calculated included area under receiver operating characteristic curve (AUROC), sensitivity, specificity, positive predictive value (PPV), and negative predictive value (NPV). The decision threshold for each model was adjusted within the test data to match the specificity of NEWS2 to facilitate comparisons of other performance metrics. The 95% confidence intervals (Cis) were calculated using bootstrap resampling with replacement (1,000 iterations). We also calculated two-sided p-values via paired permutation test with 1,000 permutations^42^. We further defined the False Alarm Ratio (FAR) of the early warning system as 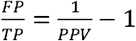, namely the number of false alarms produced for each true case detected to further evaluate the effectiveness of the model as an early warning system. Different subgroups were stratified from the held-out test set and relevant metrics were compared. For XGBoost models, feature importance was evaluated by using SHapley Additive exPlanations (SHAP) values. A TreeExplainer was applied to the trained model to compute mean absolute SHAP values for all samples in the held-out test set. These values were aggregated by features and then standardised to compare the importance of different features.

## Results

Between 01 January 2016 and 13 June 2024 there were 443,807 admissions to hospital involving 226,986 unique patients. Within this study period, we randomly sampled 3,500 time points to construct the study dataset, which based on the number of patients in the hospital at each time point resulted in 3,027,577 index date-times. There were 8,164 patients who died within 24 hours of the index date-time, the median (IQ.R) interval between the index date-time and death was 12 (6-18) hours.

Within the study dataset, 2,422,356 (80%) index date-times were used for training, 302,162 (10%) for validation (model design and tuning), and 303,059 (10%) for testing performance. The training data were down-sampled so there were equal numbers of patient index datetimes where the patient died within 24 hours and where the patient survived (n=21,814 overall). In the test data, 1,333 (0.4%) of index date-times were followed by the patient dying within 24 hours.

The median number of vital sign measurements in the 48 hours preceding each patient index date-time was 8 (IQR: 5-11) in the training set and 8 (IQR: 5-10) in validation and test sets. Most vital sign measurements fell within typical normal ranges, but had a long tailed distribution, indicating fewer cases with more severe deviations (e.g., higher heart/respiratory rates, lower oxygen saturation). There were clear differences in the distribution of vital signs between patients who survived or died within the next 24 hours for most vital signs (Figure S1).

Descriptive statistics are shown in Table 1. The median age in the training set (72 years, IQR: 63-85) was higher than the validation and test sets (67 years, IQR: 55-82, 66, IQR:54-82), due to down sampling the training set shifting the distribution. Distributions of sex, ethnicity, IMD were similar across training, validation, and test sets (Table 1).

**Table 1.** Characteristics and demographics of the training set, validation set and test set.

|  | Training* | Validation | Test |
| --- | --- | --- | --- |
| Number of observations | 21,814 | 302,162 | 303,059 |
| Age (median, IQR) (years) | 72 (63, 85) | 67 (55, 82) | 66 (54, 82) |
| Sex |  |  |  |
| *Female (%)* | *11,650 (53)* | *152,947 (51)* | *151,655 (50)* |
| *Male (%)* | *10,164 (47)* | *149,214 (49)* | *151,404 (50)* |
| Ethnicity |  |  |  |
| *White (%)* | *17,291 (79)* | *231,042 (76)* | *229,713 (76)* |
| *Mixed (%)* | *114 (1)* | *2,182 (1)* | *1,402 (0)* |
| *Asian (%)* | *403 (2)* | *5,863 (2)* | *7,219 (2)* |
| *Black (%)* | *184 (1)* | *3,699 (1)* | *4,047 (1)* |
| *Other (%)* | *180 (1)* | *2,740 (1)* | *2,933 (1)* |
| *Unknown (%)* | *3,642 (17)* | *56,635 (19)* | *57,745 (19)* |
| Charlson Comorbidity Index (median, IQR) | 3 (0, 6) | 1 (0, 5) | 1 (0, 5) |
| Index of Multiple Deprivation Score (median, IQR) | 10 (6, 16) | 11 (6, 16) | 10 (6, 16) |
| Admission Methods |  |  |  |
| *Elective (%)* | *2,305 (11)* | *56,465 (19)* | *58,655 (19)* |
| *Emergency (%)* | *19,509 (89)* | *245,696 (81)* | *244,404 (81)* |
| Number of events (%) | 10,907 (50) | 1,255 (0.4) | 1,333 (0.4) |
| Number of non-events (%) | 10,907 (50) | 300,906 (99.6) | 301,726 (99.6) |
| Number of timepoints with measured vitals in 48-hour look-back window (median, IQR) | 8 (5, 11) | 8 (5, 10) | 8 (5, 10) |

\* down-sampled to balance events

### Performance of DNNs versus comparator models using vital signs data alone

We initially investigated the performance of different predictive models using the vital signs time series alone. All models achieved an AUROC >0.90 and a sensitivity >0.7 (Figure 1, Table S3).

**Figure 1.**
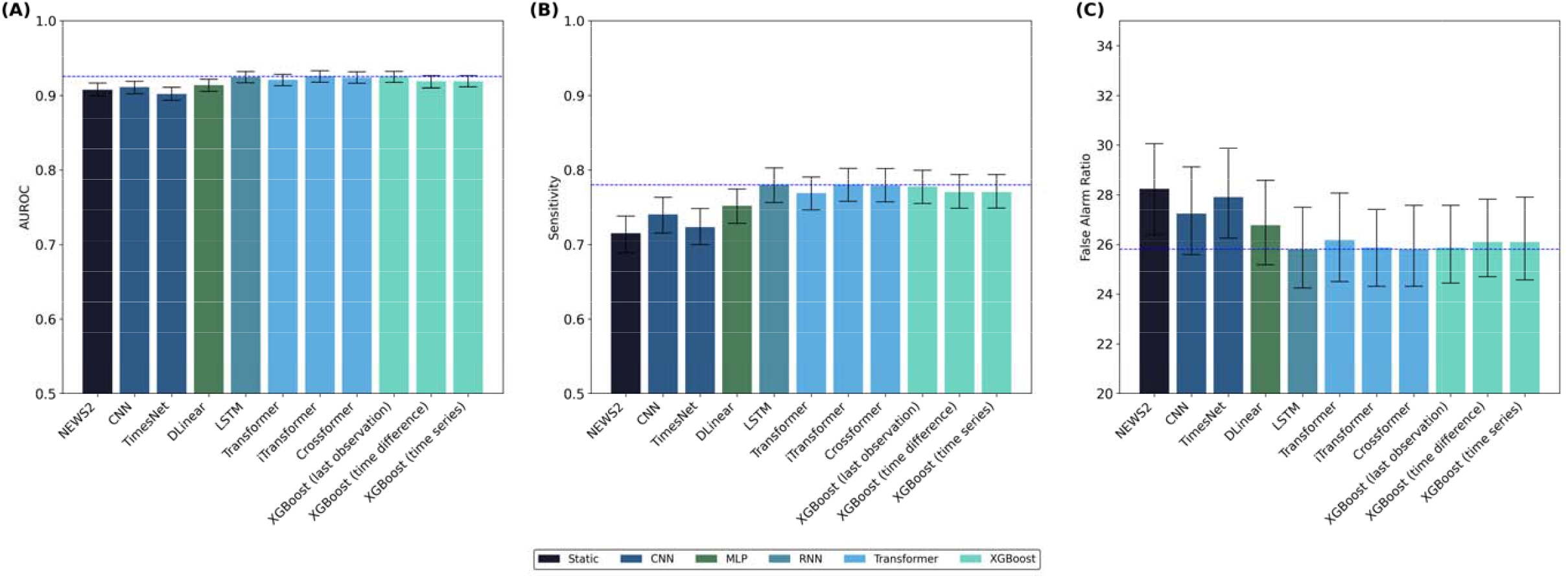
Area under the Receiver Operating Characteristic Curve (AUROC) (Panel A), Sensitivity (Panel B), and False Alarm Ratio (P different predictive models using a 48-hour lookback period. All models used data from a 48 hour look back period time series except XGBoost (last observation) and NEWS2 score that used only the last observations of vital signs. The blue dashed line indicates the best performing metric (from XGBoost with last observation) and error bars the 95% confidence intervals. The colour of the bar indicates t model. CNN, Convolutional Neural Network; LSTM, Long Short Term Memory Network.

For AUROC, all machine learning models and DNNs (regardless of architecture) outperformed NEWS2, except for TimesNet, which marginally underperformed it. All models had higher sensitivity and lower false alarm ratios than NEWS2 while maintaining (by design) the same specificity. The baseline NEWS2 model achieved an AUROC of 0.908 (95% CI: 0.899-0.916). Using only the last recorded vital signs, XGBoost achieved an AUROC of 0.925 (0.917-0.932), significantly higher than NEWS2. Incorporating more of the vital signs time series did not improve XGBoost performance. Using the last recorded vital signs and differences since the preceding measurement over a 48-hour lookback, XGBoost achieved an AUROC of 0.920 (0.912-0.927) and using the full time-series as input, XGBoost also reached an AUROC of 0.920 (0.912-0.927).

None of the DNN models significantly outperformed the classical machine learning models. Performance differences between different DNN base model types were modest, with transformer models and LSTM models having slightly higher performance compared to MLP-based models. Amongst the deep learning models. the best-performing vanilla Transformer (48-hour lookback,) achieved an AUROC of 0.921 (95% CI 0.913-0.928), LSTM achieved 0.924 (0.917-0.932), and CNN achieved 0.911 (0.902-0.919). No significant differences were observed across different variations of Transformer models, with iTransformer achieving an AUC of 0.925 (0.917-0.933) and Crossformer 0.924 (0.916-0.932).

Improvements in AUROC resulted in improvements in sensitivity compared to NEWS2 (having fixed specificity to be unchanged). Compared to NEWS2, which had a sensitivity of 0.715 (95% CI 0.689-0.738), most deep learning models achieved sensitivities between 0.76 to 0.80, with LSTM achieving 0.780 (0.756-0.803) and XGBoost only using the last observations 0.778 (0.755-0.799), an absolute increase of 6 percentage points compared to NEWS2. However, improvements in PPV were relatively modest, reflecting that these were also driven by the low prevalence of patients dying within the next 24 hours, from 0.034 (0.032-0.036) for NEWS2 to 0.037 (0.035-0.039) for XGBoost using the last observations only, and 0.037 (0.035-0.040) for the best-performing LSTM and Transformer models (Table S4).

The resulting change in false alarm ratio was from 28.2 for NEWS2 to between 26 and 27 in most models tested.

### Impact of look-back period

For models using a time series as input we investigated if using vital signs input data from different lookback windows affected model performance. Across all DNN models, the change in AUROC varying the lookback period from 12 to 48 hours was marginal, typically fluctuating within *1%* to 2%. For example, the Transformer model had an AUROCs of 0.922 (95% CI 0.914-0.930, 12 hour), 0.924 (0.916-0.932, 24 hour), and 0.921 (0.913-0.928, 48 hour), while LSTM AUROCs varied between 0.922 and 0.924 across the same lookback period. Overall, no statistically significant increase in performance was observed for any DNN models when using longer histories, and in several cases, including CNN, TimesNet, and DLinear, longer lookbacks were even associated with small declines in both AUROC and sensitivity.

In contrast, XGBoost models using time series data showed some variation with different lengths of look back periods from 2 hour to 48 hours (Figure 2). Across the whole range of lookback periods, XGBoost outperformed the NEWS2. Within the XGBoost models, the performance of time series-based approaches remained close to, but generally lower than those only using the last observation (AUROC 0.92-0.93). The best performance for the time series XGBoost models was when extending the history from very short lookback (2-4 hour) to intermediate lookback (6-24 hour); however few lookback periods outperformed the model with the last observation only even numerically, and none were significantly better. Beyond 24 hours, AUROC declined, more notably when the lookback period reached 48 hours. Similar patterns were observed for sensitivity and false alarm ratios, where the performance increased slightly with intermediate periods, and longer lookbacks were associated with a gradual drop in performance.

**Figure 2.**
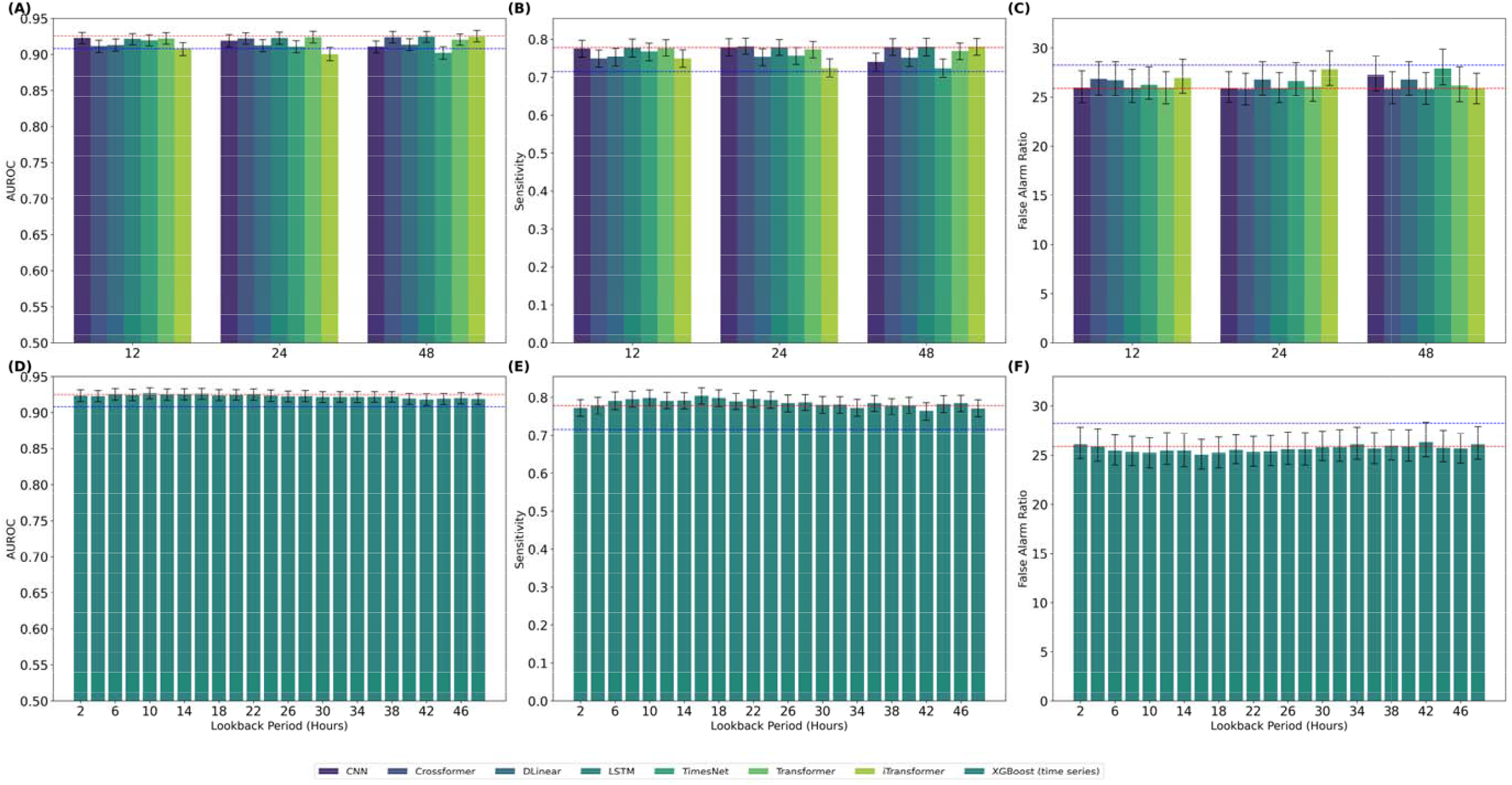
Area under the Receiver Operating Characteristic Curve (AUROC) (Panel A, D), Sensitivity (Panel B, E), and False Alarm Ra C, F) of different predictive models across lookback periods without using static information with 95% confidence interval. The err indicates the 95% confidence interval of the metric. CNN, Convolutional Neural Network; LSTM, Long Short Term Memory Network. T and red dashed line indicates the performance of NEWS2 score and XGBoost model with last observation respectively.

### Impact of adding additional static features as model inputs

We next examined whether including additional static information improved predictive performance with the same base model.

Adding static information improved the NEWS2 score AUROC from 0.908 (95% CI: 0.899-0.916) using only vital signs to 0.924 (0.916-0.930, p=0.001). This performance was comparable to many deep learning models when these used only vital sign time series as inputs. There were also improvements in the performance of XGBoost based models, e.g. adding static information improved the performance of the model using only the last vital sign observations from an AUROC of 0.925 (0.917-0.932) to 0.941 (0.936-0.948, p=0.001) with a resulting sensitivity for the new model of 0.821 (0.801-0.842), and False alarm ratio of 24.6. This was the best model overall, outperforming both deep learning models and XGBoost models using more complex time series inputs, with or without static information.

Within the DNN models, all models also showed consistently, and in most cases significantly, better performance when static features were added. For example, the AUROC of the TimesNet model with a 48-hour lookback period increased from 0.902 (95% CI 0.894-0.910) without static features to 0.921 (0.912-0.929; p=0.001) with them. The CNN model with 48-hour lookback period showed a similar pattern, improving from 0.911 (0.902-0.919) to 0.927 (0.919-0.934) when static information was added (p=0.001). The LSTM model at 48 hours exhibited a smaller AUROC gain from 0.924 (0.917-0.932) to 0.930 (0.923, 0.937), suggesting relatively limited benefit from static inputs for this architecture (p=0.006) (Figure 3).

**Figure 3.**
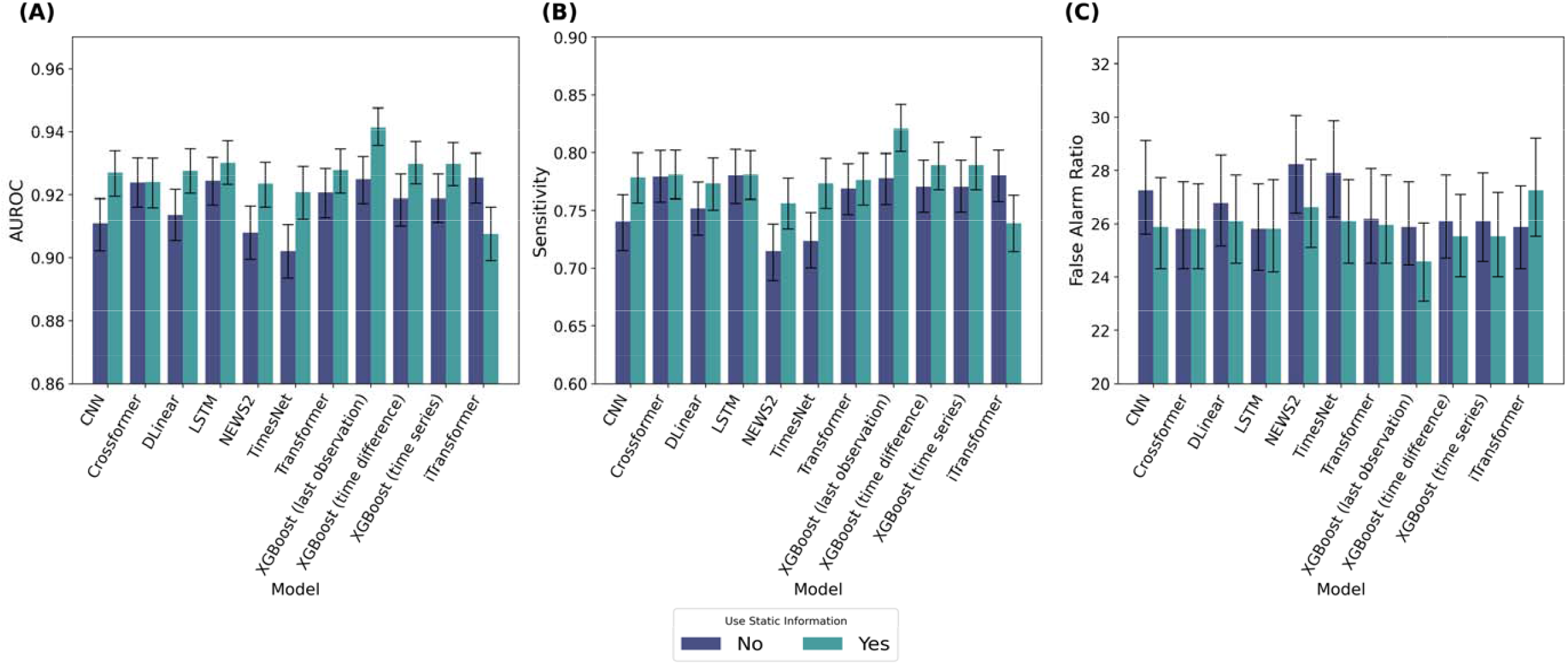
Area under the Receiver Operating Characteristic Curve (AUROC) (Panel A), Sensitivity (Panel B), and False Alarm Ratio (P different predictive models with and without usage of static information such as age, sex, comorbidities. The error bar indicates the confidence interval. CNN, Convolutional Neural Network; LSTM, Long Short Term Memory Network.

### Feature Importance Analysis

We compared feature importance values for XGBoost models with last observations only and full time series (Figure 4A). In the model with last observed values, respiratory-related vital signs had the highest SHAP importance, for example, supplemental oxygen usage had the highest importance, followed by respiratory rate and oxygen saturation. Other vital signs, such as heart rate and temperature showed lower but still important contributions.

**Figure 4.**
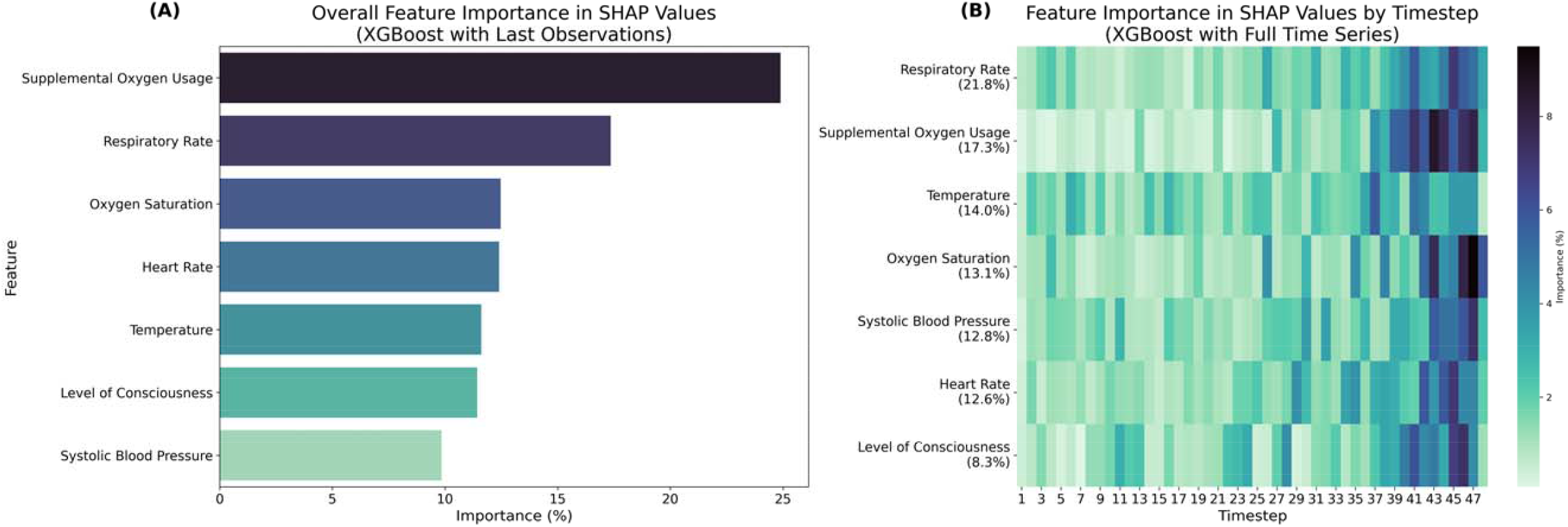
Vital signs importance in SHapley Additive exPlanations (SHAP) values in the XGBoost model with last observations only and XGBoost model with full 48-hour time series (Panel B). For both panels, features are ranked in descending order by importance color indicates greater importance of the feature in prediction.

For the model using the full time series, the aggregated importance ranking differed slightly from the model with last observations only, with the respiratory rate being the most important (21.8%), followed by supplemental oxygen usage (17.3%). Temperature (14.0%), oxygen saturation (13.1%), systolic blood pressure (12.8%), and heart rate (12.6%) contributed comparable proportions, while level of consciousness accounted for a smaller share of the total importance (8.3%). All vital signs contributed to the predictions, with no feature dominating importance. Timewise, feature importance was substantially concentrated in the timesteps immediately preceding the prediction including the last observation. For each feature, importance remained lowest and relatively uniform during starting timesteps, increasing progressively towards later timesteps (Figure 4B). Most vital signs, including supplemental oxygen, oxygen saturation, level of consciousness, exhibited a clear escalation in SHAP values at the final timestep. Few vital signs, including temperature, displayed more moderate importance, with lower peak importance but still a noticeable increase in importance as time proceeds.

### Subgroup Analysis: performance by sex, age and comorbidities

Different subgroups demonstrated moderate heterogeneity in model performance across patient characteristics between NEWS2 and the XGBoost model with the last observation (best-performing model), with XGBoost consistently showing better performance than NEWS2 in almost all examined subgroups. However, while both models had minimal differences by sex, they both experienced reduced performance in elective patients, older patients and those with severe comorbidities (Figure 5).

**Figure 5.**
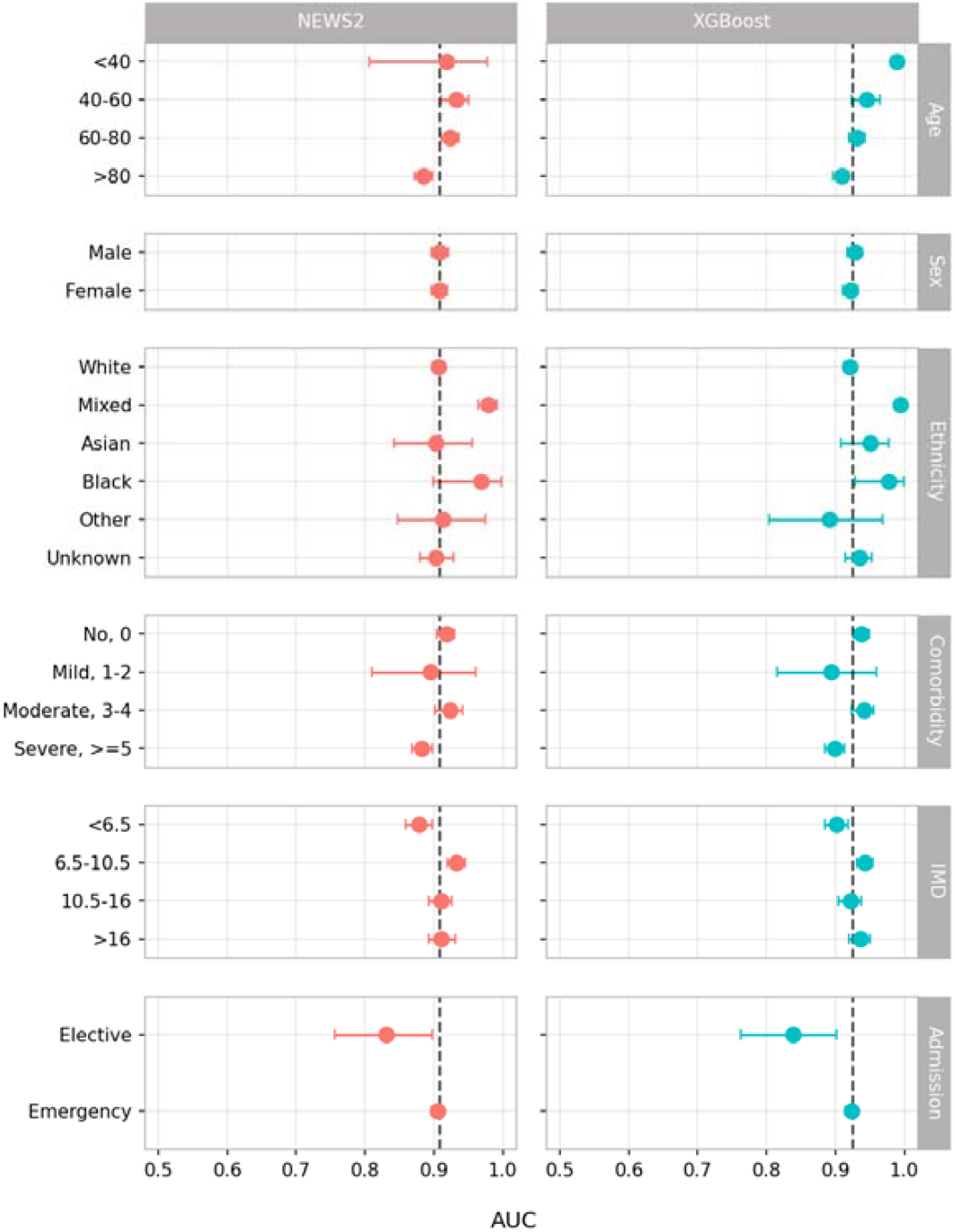
Areas under the receiver operating curve (AUC) of different patient subgroups using NEWS2 and XGBoost model using the observations only. The dashed line shows overall performance of the model.

## Discussion

In this large-scale evaluation of a wide range of models for predicting 24-hour mortality, overall, XGBoost outperformed all the deep learning models tested, even when using only the last observed vital signs rather than the whole time series. Across a range of tasks, previous research has shown that when using tabular data without time structure, XGBoost outperforms most deep learning models^29,32−34^. In our case, the data are different, and instead form a time series, where we might have expected that more data and a more sophisticated model would improve performance, for example by identifying patients who are deteriorating or improving, as well as using the absolute values of their vital signs. However, XGBoost using only the last vital sign observations performed better than models where the input of a neural network is formatted as a time series. This is surprising in the context of medical teaching that often focuses on identifying patient deterioration by looking at changes over time. In fact, here we find, at least for mortality prediction, it may be more important ‘where you end up’ rather than ‘how you got there’ when analysing vital signs, and that the last observation is typically the most informative. This finding applies to mortality prediction over the next 24 hours, with time-to-death being approximately uniformly distributed over the 24 hour period, i.e., it is not the case we are simply predicting imminent death from very abnormal vital signs just before. XGBoost also offered performance improvements in different patient subgroups over a simpler scoring system, NEWS2, albeit with an increase in model complexity and potentially some reduction in transparency for clinicians in how predictions are calculated (which could be addressed). Adding information on demographics and comorbidities improved model performance further.

Respiratory-related variables showed substantial importance in predicting in-hospital mortality, particularly supplemental oxygen usage and respiratory rate, which together accounted for a considerable proportion of the total feature importance in the XGBoost models. When using time series, SHAP values for all features were consistently highest in the final timesteps: earlier measurements contributed comparatively little. This concentration of importance near the prediction time indicated that, even when historical data are available, the model might primarily rely on recent physiological measurements. This reflects that the effective predictive signal contained in the full time series might have largely overlapped with that captured in the last observation. Therefore, the marginal benefit of incorporating historical context might not outweigh the added noise and model complexity relative to a snapshot formulation.

From the perspective of deployment, XGBoost has significant advantages in computational efficiency compared to deep learning models, including the lower training and inference times, as well as reduced demand of resources, labour and financial cost^33,34^. XGBoost models are also naturally more interpretable as a tree-based model^33^, which makes the assessment of outputs by healthcare professionals more tractable and has more established cases in real-world application^43^. Other baseline models, such as NEWS2, also achieved competitive performance using a linear scoring model. Such a scoring system provides clear clinical instructions and allows the healthcare professionals to understand the output^28^.

Fortunately, inpatient mortality is a rare outcome. This means that even with improvements in model performance false alarm rates remained higher than ideal. For example, comparing NEWS2 to the best performing XGBoost model that also included static data as inputs reduced the false alarm ratio from 28.2 false alarms per true alarm to 24.6, meaning that only approximately 3 false alarms would be prevented per true alarm by changing model. However, ‘false alarms’ on NEWS2 may have triggered an intervention that prevented a death, and so they are not necessarily unhelpful. Since deploying a machine learning model involves additional cost in various sources such as computation, hardware updates and staff training, the further benefit of deploying machine learning models as an early warning system with only this modest improvement remains to be investigated.

None of the DNN models tested offered better performance than XGBoost, despite testing a wide range of architectures and a range of hyperparameters for each model. Given the trend towards DNN models being widely accepted and prioritised by the research community, we encourage that when comparing different models for inpatient care, existing tools like the NEWS2 and tree-based methods such as XGBoost, and non-time series features like demographics and comorbidities, should also be considered carefully since it may be possible to achieve optimal performance by using more established methods that are also potentially more interpretable and lower cost.

Our study has several limitations. We cannot rule out the possibility that more sophisticated deep learning could better predict risks, including applications of large language models (LLM) and foundation models to time series^44−46^. Additionally, we also only considered a limited number of vital signs instead of making use of other features in patient records such as laboratory and imaging results and consultation notes. Incorporating these features might further leverage the advantages of deep learning when processing larger and higher-dimensional data. It is also possible that time series models would perform better where vital signs are measured continuously or more frequently, such as ICU environments; in this study the median number of sets of vital signs recorded within 48 hour was 8, with this relatively sparse data requiring other hourly timesteps to be interpolated. However, this frequency of measurement is typical for many non-ICU inpatient settings where early warning systems are most frequently used. We did not conduct any external validation for this study: however data from 4 hospitals covering a wide range of specialities and populations were included.

## Conclusion

When predicting 24-hour mortality in inpatient care, multiple different DNN architectures using vital signs time series as inputs did not yield better performance than an XGBoost model using only the last observation of the time series. Predictive performance of XGBoost models was better than a clinical score, NEWS2, especially when also considering demographic data and comorbidities. However, reductions in false alarms implementing this approach would only be modest. Careful evaluation of established approaches should be included alongside applications of newer methods in studies developing predictive healthcare models.

## Data Availability

The datasets analysed during the current study are not publicly available as they contain personal data but are available from the Infections in Oxfordshire Research Database (https://oxfordbrc.nihr.ac.uk/research-themes/modernising-medical-microbiology-and-big-infection-diagnostics/iord-about/), subject to an application and research proposal meeting the ethical and governance requirements of the Database. For further details on how to apply for access to the data and a research proposal template please.

## Author Contributions

The study was designed and conceived by ZZ and DWE. ZZ, JW and DWE curated the data. ZZ, JX, YL, HC, SB, and AL analysed the data and created visualisations. ZZ, JW, and DWE wrote the first draft of the manuscript. ASW, DAC and all other authors contributed to editing and revising the manuscript.

## Code Availability

All analysis code can be accessed via the following link: https://github.com/zizhengzhang1/less_is_more_benchmark.

## Ethics Committee Approval

Deidentified data covering patient demographics, mortality, hospital admissions, diagnostic codes and vital signs were obtained from the Infections in Oxfordshire Research Database, which has approvals from the National Research Ethics Service South Central-Oxford C Research Ethics Committee (24/SC/0241), Health Research Authority and Confidentiality Advisory Group (19/CAG/0144) as a deidentified database without individual consent.

## Acknowledgements

This project was funded by the National Institute for Health and Care Research (NIHR) Health Protection Research Unit in Healthcare Associated Infections and Antimicrobial Resistance at Oxford University in partnership with the UK Health Security Agency (UKHSA) (NIHR207397) and the NIHR Biomedical Research Centre, Oxford.

The views expressed are those of the authors and not necessarily those of the NHS, the NIHR, the Department of Health and Social Care or UKHSA. ASW is an NIHR Senior Investigator.

For the purpose of Open Access, the author has applied a CC BY public copyright licence to any Author Accepted Manuscript (AAM) version arising.

This work uses data provided by patients and collected by the UK’s National Health Service as part of their care and support. We thank all the people of Oxfordshire who contribute to the Infections in Oxfordshire Research Database.

Research Database Team: L Butcher, H Boseley, C Crichton, DW Crook, J Davies, D Eyre, R Harrington, G Hayward, K Jeffery, F Kemp, E Morris, TEA Peto, D Prieto-Alhambra, TP Quan, R Shackell, B Shine, AS Walker, K Woods.

Patient and Public Panel: M Ahmed, G Blower, J Hopkins, R Mandunya, S Markham.

