## Supplemental material for "Less is More: last observations of vital signs can outperform time series for hospital mortality prediction"

### A.1 Additional Pre-processing Details of Features

**Table S1. The upper and lower limit of vital signs recommended by the clinicians.**

| **Metric** | **Lower limit** | **Upper limit** |
| --- | --- | --- |
| Heart rate, beats per minute | 30 | 300 |
| Respiratory rate, breaths per minute | 5 | 80 |
| Temperature, degrees Celcius | 25 | 45 |
| Systolic blood pressure, mmHg | 60 | 250 |
| Oxygen saturation, % | 70 | 100 |
| Use of supplemental oxygen, litres per minute | 0 | 100 |
| Level of consciousness, see footnote | 0 | 3 |

As different scales were used to record the level of consciousness, a combined feature was created, derived from Glasgow Coma Score (GCS), AVPU scale (alert, verbal, pain, unresponsive) and ACVPU scale (alert, confusion, verbal, pain, unresponsive), depending on which score was recorded for the patient. The combined feature was an ordinal variable with 4 classes, where class 0 corresponded to ‘Alert’ in AVPU and ACVPU and 15 in GCS; class 1 corresponded to ‘Verbal’ in AVPU, ‘Verbal’ and ‘Confusion’ in ACVPU and 11-14 in GCS; class 2 corresponded to ‘Pain’ in AVPU and ACVPU and 4-10 in GCS; class 3 corresponded to ‘Unresponsive’ in AVPU and ACVPU and 3 in GCS.

**Table S2. The details of the static features used**

| **Feature** | **Type of Feature** | **Feature Description** |
| --- | --- | --- |
| Sex | Categorical | Contains 2 categories, i.e. male/ female |
| Age | Numerical | Rounded into years |
| Ethnicity | Categorical | 6 categories, namely White, Asian, Black, Other, Mixed, Unknown |
| Comorbidities | Numerical | Charlson comorbidity index calculated using ICD 10 codes from admissions in the prior 1 year |
| Index of Multiple Deprivation score | Numerical | Numerical feature measuring deprivation of each patient’s residential area |
| Admission method | Categorical | Contains 2 categories, i.e. emergency and elective |

### A.2 Distribution of Patient Vital Signs

**Figure S1. Distribution of Patient Vital Signs in the Study Dataset (i.e., training, validation and test datasets combined).**

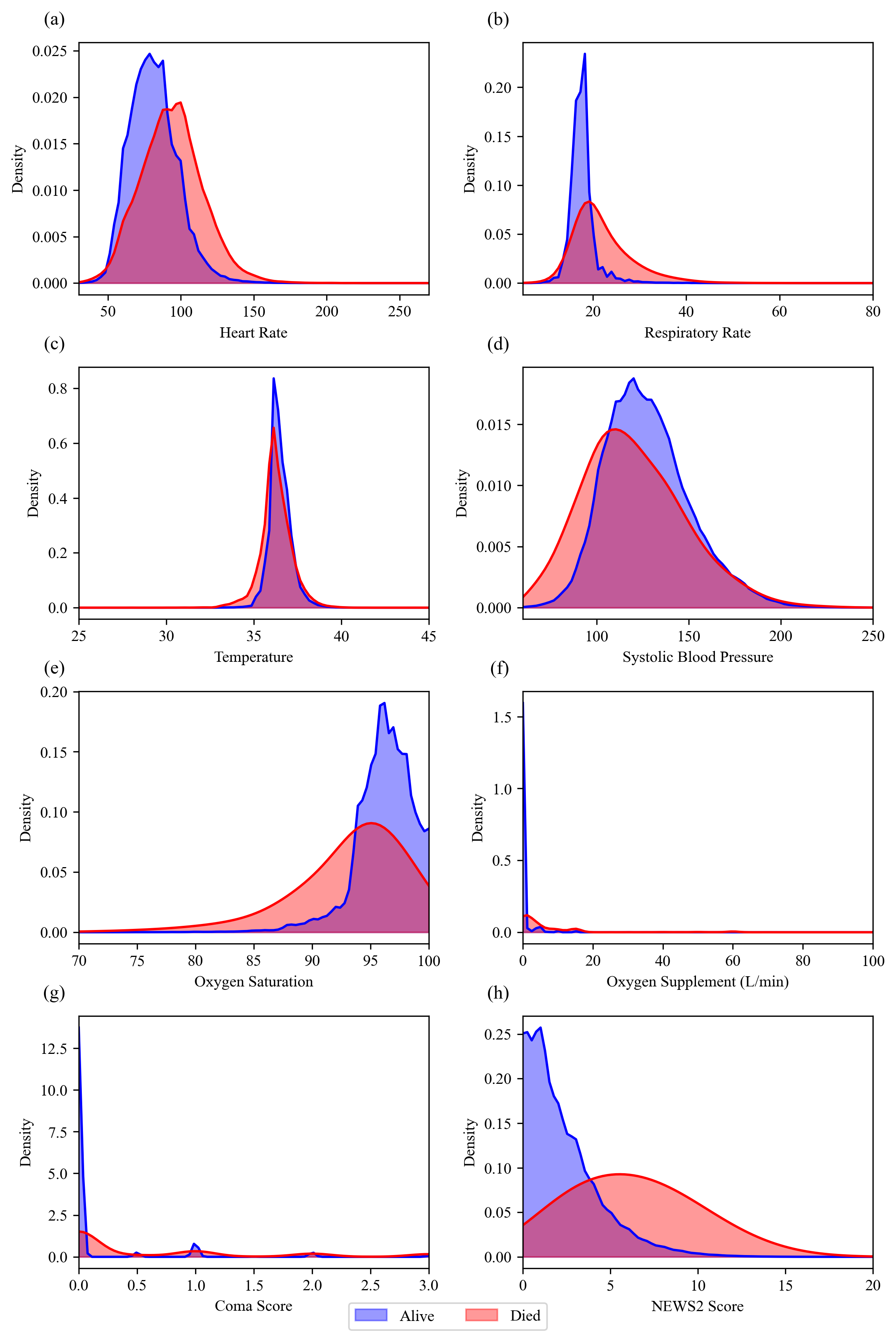

### A.3 Training Configurations

The batch size of the input time series was set to 64 and the initial learning rate was set as 0.001 with linear decay. Models were trained with 30 epochs with early stopping mechanisms. We used binary cross entropy as the loss function and Adam as the optimiser.

The hyperparameter searching range is given as follows.

- Convolutional Neural Network (CNN): The CNN consists of 2 convolutional layers. The kernel size is in {2, 4, 6} for both layers, and the out channels size is in {16, 32, 64, 128} and {8, 16, 32}. The weight decay of the hyperparameter is in {0, 0.001, 0.00001}.
- TimesNet: The number of embedding layers is in {1, 2}. The depth of the model is given by {4, 8, 16}. The number of heads is given by {1, 2}. The dimension of feedforward is in {4, 8}. The number of top frequencies is {1, 2}. The number of convolutional kernels is given by {2, 4}. The dropout rate is in {0, 0.2, 0.5}. The weight decay of the hyperparameter is in {0, 0.001, 0.00001}.
- Long Short Term Memory networks (LSTM): The number of LSTM layers is in {1, 2}. The hidden size of the LSTM unit is in {8, 16, 32, 64}. The dropout rate is in {0, 0.3, 0.5}. The weight decay of the hyperparameter is in {0, 0.001, 0.00001}.
- DLinear: The length of moving average window is in {5, 15, 25}. The weight decay of the hyperparameter is in {0, 0.001, 0.00001}.
- Vanilla Transformer, iTransformer, crossformer: The number of embedding layers is in {1, 2}. The depth of the model is given by {8, 16}. The learning factor is in {2, 4}. The number of heads is given by {2, 4}. The dimension of feedforward is in {8, 16}. The dropout rate is in {0, 0.2, 0.5}. The weight decay of the hyperparameter is in {0, 0.001, 0.00001}.

### A.4 Full results on model performance

**Table S3. The performance of all models (AUROC, sensitivity, specificity). AUROC stands for Area under the Receiver Operating Characteristic Curve, CNN stands for Convolutional Neural Network, LSTM stands for Long Short Term Memory Network**

| Model | Lookback (hours) | Interpolation | Static | AUROC | Sensitivity | Specificity |
| --- | --- | --- | --- | --- | --- | --- |
| CNN | 12 | Linear | 0 | 0.923 (0.915, 0.930) | 0.776 (0.753, 0.797) | 0.911 (0.910, 0.912) |
| CNN | 12 | Linear | 1 | 0.933 (0.926, 0.939) | 0.793 (0.771, 0.814) | 0.911 (0.910, 0.912) |
| CNN | 24 | Linear | 0 | 0.919 (0.910, 0.927) | 0.779 (0.756, 0.802) | 0.911 (0.910, 0.912) |
| CNN | 24 | Linear | 1 | 0.933 (0.926, 0.939) | 0.803 (0.781, 0.822) | 0.911 (0.910, 0.912) |
| CNN | 48 | Linear | 0 | 0.911 (0.902, 0.919) | 0.740 (0.715, 0.763) | 0.911 (0.910, 0.912) |
| CNN | 48 | Linear | 1 | 0.927 (0.919, 0.934) | 0.779 (0.756, 0.800) | 0.911 (0.910, 0.912) |
| Crossformer | 12 | Linear | 0 | 0.912 (0.903, 0.920) | 0.749 (0.726, 0.772) | 0.911 (0.910, 0.912) |
| Crossformer | 12 | Linear | 1 | 0.924 (0.916, 0.931) | 0.776 (0.755, 0.797) | 0.911 (0.910, 0.912) |
| Crossformer | 24 | Linear | 0 | 0.922 (0.914, 0.930) | 0.781 (0.761, 0.803) | 0.911 (0.910, 0.912) |
| Crossformer | 24 | Linear | 1 | 0.920 (0.912, 0.928) | 0.770 (0.748, 0.792) | 0.911 (0.910, 0.912) |
| Crossformer | 48 | Linear | 0 | 0.924 (0.916, 0.932) | 0.779 (0.757, 0.802) | 0.911 (0.910, 0.912) |
| Crossformer | 48 | Linear | 1 | 0.924 (0.916, 0.932) | 0.781 (0.760, 0.802) | 0.911 (0.910, 0.912) |
| DLinear | 12 | Linear | 0 | 0.913 (0.904, 0.921) | 0.754 (0.730, 0.776) | 0.911 (0.910, 0.912) |
| DLinear | 12 | Linear | 1 | 0.928 (0.922, 0.935) | 0.772 (0.749, 0.794) | 0.911 (0.910, 0.912) |
| DLinear | 24 | Linear | 0 | 0.912 (0.904, 0.921) | 0.753 (0.730, 0.775) | 0.911 (0.910, 0.912) |
| DLinear | 24 | Linear | 1 | 0.927 (0.920, 0.934) | 0.776 (0.754, 0.797) | 0.911 (0.910, 0.912) |
| DLinear | 48 | Linear | 0 | 0.914 (0.905, 0.922) | 0.752 (0.728, 0.774) | 0.911 (0.910, 0.912) |
| DLinear | 48 | Linear | 1 | 0.928 (0.920, 0.934) | 0.773 (0.750, 0.796) | 0.911 (0.910, 0.912) |
| LSTM | 12 | Linear | 0 | 0.922 (0.913, 0.929) | 0.777 (0.753, 0.801) | 0.911 (0.910, 0.912) |
| LSTM | 12 | Linear | 1 | 0.930 (0.923, 0.936) | 0.772 (0.750, 0.795) | 0.911 (0.910, 0.912) |
| LSTM | 24 | Linear | 0 | 0.923 (0.914, 0.931) | 0.779 (0.757, 0.799) | 0.911 (0.910, 0.912) |
| LSTM | 24 | Linear | 1 | 0.932 (0.925, 0.939) | 0.797 (0.775, 0.817) | 0.911 (0.910, 0.912) |
| LSTM | 48 | Linear | 0 | 0.924 (0.917, 0.932) | 0.780 (0.756, 0.803) | 0.911 (0.910, 0.912) |
| LSTM | 48 | Linear | 1 | 0.930 (0.923, 0.937) | 0.781 (0.759, 0.801) | 0.911 (0.910, 0.912) |
| NEWS2 |  |  | 0 | 0.908 (0.899, 0.916) | 0.715 (0.689, 0.738) | 0.911 (0.910, 0.912) |
| NEWS2 |  |  | 1 | 0.924 (0.916, 0.930) | 0.756 (0.734, 0.778) | 0.911 (0.910, 0.912) |
| TimesNet | 12 | Linear | 0 | 0.920 (0.912, 0.927) | 0.767 (0.743, 0.790) | 0.911 (0.910, 0.912) |
| TimesNet | 12 | Linear | 1 | 0.921 (0.913, 0.928) | 0.770 (0.746, 0.793) | 0.911 (0.910, 0.912) |
| TimesNet | 24 | Linear | 0 | 0.911 (0.902, 0.919) | 0.756 (0.734, 0.778) | 0.911 (0.910, 0.912) |
| TimesNet | 24 | Linear | 1 | 0.914 (0.906, 0.921) | 0.744 (0.720, 0.767) | 0.911 (0.910, 0.912) |
| TimesNet | 48 | Linear | 0 | 0.902 (0.894, 0.910) | 0.723 (0.700, 0.748) | 0.911 (0.910, 0.912) |
| TimesNet | 48 | Linear | 1 | 0.921 (0.912, 0.929) | 0.773 (0.752, 0.795) | 0.911 (0.910, 0.912) |
| Transformer | 12 | Linear | 0 | 0.922 (0.914, 0.930) | 0.776 (0.755, 0.799) | 0.911 (0.910, 0.912) |
| Transformer | 12 | Linear | 1 | 0.932 (0.925, 0.939) | 0.780 (0.757, 0.802) | 0.911 (0.910, 0.912) |
| Transformer | 24 | Linear | 0 | 0.924 (0.916, 0.932) | 0.773 (0.751, 0.794) | 0.911 (0.910, 0.912) |
| Transformer | 24 | Linear | 1 | 0.932 (0.925, 0.938) | 0.794 (0.773, 0.816) | 0.911 (0.910, 0.912) |
| Transformer | 48 | Linear | 0 | 0.921 (0.913, 0.928) | 0.769 (0.746, 0.790) | 0.911 (0.910, 0.912) |
| Transformer | 48 | Linear | 1 | 0.928 (0.920, 0.934) | 0.776 (0.755, 0.799) | 0.911 (0.910, 0.912) |
| XGBoost (last observation) |  |  | 0 | 0.925 (0.917, 0.932) | 0.778 (0.755, 0.799) | 0.911 (0.910, 0.912) |
| XGBoost (last observation) |  |  | 1 | 0.941 (0.936, 0.948) | 0.821 (0.801, 0.842) | 0.911 (0.910, 0.912) |
| XGBoost (time difference) | 12 |  | 0 | 0.925 (0.917, 0.933) | 0.791 (0.771, 0.813) | 0.911 (0.910, 0.912) |
| XGBoost (time difference) | 12 |  | 1 | 0.940 (0.933, 0.946) | 0.822 (0.802, 0.843) | 0.911 (0.910, 0.912) |
| XGBoost (time difference) | 24 |  | 0 | 0.924 (0.916, 0.932) | 0.793 (0.771, 0.814) | 0.911 (0.910, 0.912) |
| XGBoost (time difference) | 24 |  | 1 | 0.937 (0.930, 0.943) | 0.817 (0.797, 0.837) | 0.911 (0.910, 0.912) |
| XGBoost (time difference) | 48 |  | 0 | 0.919 (0.910, 0.926) | 0.770 (0.748, 0.794) | 0.911 (0.910, 0.912) |
| XGBoost (time difference) | 48 |  | 1 | 0.930 (0.923, 0.937) | 0.789 (0.768, 0.809) | 0.911 (0.910, 0.912) |
| XGBoost (time series) | 2 |  | 0 | 0.924 (0.915, 0.932) | 0.772 (0.750, 0.794) | 0.911 (0.910, 0.912) |
| XGBoost (time series) | 4 |  | 0 | 0.923 (0.915, 0.931) | 0.778 (0.756, 0.800) | 0.911 (0.910, 0.912) |
| XGBoost (time series) | 6 |  | 0 | 0.926 (0.917, 0.933) | 0.791 (0.767, 0.814) | 0.911 (0.910, 0.912) |
| XGBoost (time series) | 8 |  | 0 | 0.924 (0.916, 0.932) | 0.795 (0.774, 0.816) | 0.911 (0.910, 0.912) |
| XGBoost (time series) | 10 |  | 0 | 0.927 (0.919, 0.934) | 0.798 (0.778, 0.820) | 0.911 (0.910, 0.912) |
| XGBoost (time series) | 12 |  | 0 | 0.925 (0.917, 0.933) | 0.791 (0.770, 0.813) | 0.911 (0.910, 0.912) |
| XGBoost (time series) | 12 |  | 1 | 0.940 (0.934, 0.945) | 0.822 (0.802, 0.843) | 0.911 (0.910, 0.912) |
| XGBoost (time series) | 14 |  | 0 | 0.926 (0.917, 0.933) | 0.791 (0.769, 0.812) | 0.911 (0.910, 0.912) |
| XGBoost (time series) | 16 |  | 0 | 0.926 (0.918, 0.933) | 0.804 (0.782, 0.825) | 0.911 (0.910, 0.912) |
| XGBoost (time series) | 18 |  | 0 | 0.924 (0.916, 0.932) | 0.798 (0.775, 0.820) | 0.911 (0.910, 0.912) |
| XGBoost (time series) | 20 |  | 0 | 0.924 (0.917, 0.932) | 0.789 (0.768, 0.810) | 0.911 (0.910, 0.912) |
| XGBoost (time series) | 22 |  | 0 | 0.925 (0.917, 0.933) | 0.796 (0.774, 0.818) | 0.911 (0.910, 0.912) |
| XGBoost (time series) | 24 |  | 0 | 0.924 (0.915, 0.931) | 0.793 (0.772, 0.815) | 0.911 (0.910, 0.912) |
| XGBoost (time series) | 24 |  | 1 | 0.937 (0.930, 0.943) | 0.817 (0.795, 0.837) | 0.911 (0.910, 0.912) |
| XGBoost (time series) | 26 |  | 0 | 0.923 (0.915, 0.930) | 0.785 (0.761, 0.806) | 0.911 (0.910, 0.912) |
| XGBoost (time series) | 28 |  | 0 | 0.923 (0.915, 0.930) | 0.787 (0.765, 0.807) | 0.911 (0.910, 0.912) |
| XGBoost (time series) | 30 |  | 0 | 0.922 (0.914, 0.929) | 0.780 (0.757, 0.802) | 0.911 (0.910, 0.912) |
| XGBoost (time series) | 32 |  | 0 | 0.922 (0.914, 0.929) | 0.781 (0.758, 0.802) | 0.911 (0.910, 0.912) |
| XGBoost (time series) | 34 |  | 0 | 0.922 (0.914, 0.929) | 0.772 (0.750, 0.795) | 0.911 (0.910, 0.912) |
| XGBoost (time series) | 36 |  | 0 | 0.922 (0.914, 0.929) | 0.785 (0.762, 0.805) | 0.911 (0.910, 0.912) |
| XGBoost (time series) | 38 |  | 0 | 0.922 (0.915, 0.929) | 0.777 (0.755, 0.797) | 0.911 (0.910, 0.912) |
| XGBoost (time series) | 40 |  | 0 | 0.919 (0.911, 0.926) | 0.779 (0.756, 0.800) | 0.911 (0.910, 0.912) |
| XGBoost (time series) | 42 |  | 0 | 0.918 (0.910, 0.926) | 0.764 (0.740, 0.786) | 0.911 (0.910, 0.912) |
| XGBoost (time series) | 44 |  | 0 | 0.919 (0.911, 0.926) | 0.782 (0.759, 0.804) | 0.911 (0.910, 0.912) |
| XGBoost (time series) | 46 |  | 0 | 0.920 (0.912, 0.927) | 0.785 (0.762, 0.806) | 0.911 (0.910, 0.912) |
| XGBoost (time series) | 48 |  | 0 | 0.919 (0.911, 0.926) | 0.770 (0.748, 0.794) | 0.911 (0.910, 0.912) |
| XGBoost (time series) | 48 |  | 1 | 0.930 (0.923, 0.936) | 0.789 (0.768, 0.813) | 0.911 (0.910, 0.912) |
| iTransformer | 12 | Linear | 0 | 0.908 (0.898, 0.916) | 0.749 (0.726, 0.772) | 0.911 (0.910, 0.912) |
| iTransformer | 12 | Linear | 1 | 0.915 (0.907, 0.924) | 0.764 (0.742, 0.787) | 0.911 (0.910, 0.912) |
| iTransformer | 24 | Linear | 0 | 0.900 (0.891, 0.910) | 0.724 (0.701, 0.748) | 0.911 (0.910, 0.912) |
| iTransformer | 24 | Linear | 1 | 0.917 (0.908, 0.925) | 0.764 (0.741, 0.786) | 0.911 (0.910, 0.912) |
| iTransformer | 48 | Linear | 0 | 0.925 (0.917, 0.933) | 0.780 (0.758, 0.802) | 0.911 (0.910, 0.912) |
| iTransformer | 48 | Linear | 1 | 0.908 (0.899, 0.916) | 0.739 (0.714, 0.763) | 0.911 (0.910, 0.912) |

**Table S4. The performance (95% Confidence Intervals) of all models (PPV, NPV, FAR). PPV stands for Positive Predictive Value, NPV stands for Negative Predictive Value, FAR stands for False Alarm Ratio. CNN stands for Convolutional Neural Network, LSTM stands for Long Short Term Memory Network**

| Model | Lookback | Interpolation | Static | PPV | NPV | FAR |
| --- | --- | --- | --- | --- | --- | --- |
| CNN | 12 | Linear | 0 | 0.037 (0.035, 0.039) | 0.999 (0.999, 0.999) | 25.954 (24.445, 27.653) |
| CNN | 12 | Linear | 1 | 0.038 (0.036, 0.040) | 0.999 (0.999, 0.999) | 25.385 (23.938, 26.933) |
| CNN | 24 | Linear | 0 | 0.037 (0.035, 0.039) | 0.999 (0.999, 0.999) | 25.882 (24.445, 27.571) |
| CNN | 24 | Linear | 1 | 0.038 (0.036, 0.040) | 0.999 (0.999, 0.999) | 25.110 (23.691, 26.624) |
| CNN | 48 | Linear | 0 | 0.035 (0.033, 0.038) | 0.999 (0.999, 0.999) | 27.249 (25.596, 29.120) |
| CNN | 48 | Linear | 1 | 0.037 (0.035, 0.040) | 0.999 (0.999, 0.999) | 25.882 (24.316, 27.736) |
| Crossformer | 12 | Linear | 0 | 0.036 (0.034, 0.038) | 0.999 (0.999, 0.999) | 26.855 (25.178, 28.586) |
| Crossformer | 12 | Linear | 1 | 0.037 (0.035, 0.039) | 0.999 (0.999, 0.999) | 25.954 (24.445, 27.736) |
| Crossformer | 24 | Linear | 0 | 0.037 (0.035, 0.040) | 0.999 (0.999, 0.999) | 25.810 (24.189, 27.409) |
| Crossformer | 24 | Linear | 1 | 0.037 (0.035, 0.039) | 0.999 (0.999, 0.999) | 26.174 (24.575, 27.902) |
| Crossformer | 48 | Linear | 0 | 0.037 (0.035, 0.040) | 0.999 (0.999, 0.999) | 25.810 (24.316, 27.571) |
| Crossformer | 48 | Linear | 1 | 0.037 (0.035, 0.040) | 0.999 (0.999, 0.999) | 25.810 (24.316, 27.490) |
| DLinear | 12 | Linear | 0 | 0.036 (0.034, 0.038) | 0.999 (0.999, 0.999) | 26.701 (25.110, 28.586) |
| DLinear | 12 | Linear | 1 | 0.037 (0.035, 0.039) | 0.999 (0.999, 0.999) | 26.100 (24.510, 27.736) |
| DLinear | 24 | Linear | 0 | 0.036 (0.034, 0.038) | 0.999 (0.999, 0.999) | 26.778 (25.178, 28.586) |
| DLinear | 24 | Linear | 1 | 0.037 (0.035, 0.039) | 0.999 (0.999, 0.999) | 25.954 (24.381, 27.653) |
| DLinear | 48 | Linear | 0 | 0.036 (0.034, 0.038) | 0.999 (0.999, 0.999) | 26.778 (25.178, 28.586) |
| DLinear | 48 | Linear | 1 | 0.037 (0.035, 0.039) | 0.999 (0.999, 0.999) | 26.100 (24.510, 27.818) |
| LSTM | 12 | Linear | 0 | 0.037 (0.035, 0.039) | 0.999 (0.999, 0.999) | 25.954 (24.445, 27.818) |
| LSTM | 12 | Linear | 1 | 0.037 (0.035, 0.039) | 0.999 (0.999, 0.999) | 26.100 (24.510, 27.818) |
| LSTM | 24 | Linear | 0 | 0.037 (0.035, 0.039) | 0.999 (0.999, 0.999) | 25.882 (24.445, 27.490) |
| LSTM | 24 | Linear | 1 | 0.038 (0.036, 0.040) | 0.999 (0.999, 0.999) | 25.316 (23.752, 27.090) |
| LSTM | 48 | Linear | 0 | 0.037 (0.035, 0.040) | 0.999 (0.999, 0.999) | 25.810 (24.253, 27.490) |
| LSTM | 48 | Linear | 1 | 0.037 (0.035, 0.040) | 0.999 (0.999, 0.999) | 25.810 (24.189, 27.653) |
| NEWS2 |  |  | 0 | 0.034 (0.032, 0.036) | 0.999 (0.998, 0.999) | 28.240 (26.397, 30.056) |
| NEWS2 |  |  | 1 | 0.036 (0.034, 0.038) | 0.999 (0.999, 0.999) | 26.624 (25.110, 28.412) |
| TimesNet | 12 | Linear | 0 | 0.037 (0.034, 0.039) | 0.999 (0.999, 0.999) | 26.248 (24.773, 28.070) |
| TimesNet | 12 | Linear | 1 | 0.037 (0.035, 0.039) | 0.999 (0.999, 0.999) | 26.174 (24.641, 27.818) |
| TimesNet | 24 | Linear | 0 | 0.036 (0.034, 0.038) | 0.999 (0.999, 0.999) | 26.624 (25.110, 28.499) |
| TimesNet | 24 | Linear | 1 | 0.036 (0.033, 0.038) | 0.999 (0.999, 0.999) | 27.090 (25.455, 28.940) |
| TimesNet | 48 | Linear | 0 | 0.035 (0.032, 0.037) | 0.999 (0.998, 0.999) | 27.902 (26.248, 29.864) |
| TimesNet | 48 | Linear | 1 | 0.037 (0.035, 0.039) | 0.999 (0.999, 0.999) | 26.100 (24.510, 27.653) |
| Transformer | 12 | Linear | 0 | 0.037 (0.035, 0.040) | 0.999 (0.999, 0.999) | 25.954 (24.316, 27.571) |
| Transformer | 12 | Linear | 1 | 0.037 (0.035, 0.040) | 0.999 (0.999, 0.999) | 25.810 (24.189, 27.653) |
| Transformer | 24 | Linear | 0 | 0.037 (0.035, 0.039) | 0.999 (0.999, 0.999) | 26.027 (24.575, 27.653) |
| Transformer | 24 | Linear | 1 | 0.038 (0.036, 0.040) | 0.999 (0.999, 0.999) | 25.385 (23.876, 27.011) |
| Transformer | 48 | Linear | 0 | 0.037 (0.034, 0.039) | 0.999 (0.999, 0.999) | 26.174 (24.510, 28.070) |
| Transformer | 48 | Linear | 1 | 0.037 (0.035, 0.039) | 0.999 (0.999, 0.999) | 25.954 (24.510, 27.818) |
| XGBoost (last observation) |  |  | 0 | 0.037 (0.035, 0.039) | 0.999 (0.999, 0.999) | 25.882 (24.445, 27.571) |
| XGBoost (last observation) |  |  | 1 | 0.039 (0.037, 0.042) | 0.999 (0.999, 0.999) | 24.575 (23.096, 26.027) |
| XGBoost (time difference) | 12 |  | 0 | 0.038 (0.036, 0.040) | 0.999 (0.999, 0.999) | 25.455 (24.063, 27.090) |
| XGBoost (time difference) | 12 |  | 1 | 0.039 (0.037, 0.042) | 0.999 (0.999, 0.999) | 24.510 (23.096, 26.100) |
| XGBoost (time difference) | 24 |  | 0 | 0.038 (0.036, 0.040) | 0.999 (0.999, 0.999) | 25.385 (23.938, 27.011) |
| XGBoost (time difference) | 24 |  | 1 | 0.039 (0.037, 0.041) | 0.999 (0.999, 0.999) | 24.641 (23.213, 26.322) |
| XGBoost (time difference) | 48 |  | 0 | 0.037 (0.035, 0.039) | 0.999 (0.999, 0.999) | 26.100 (24.707, 27.818) |
| XGBoost (time difference) | 48 |  | 1 | 0.038 (0.036, 0.040) | 0.999 (0.999, 0.999) | 25.525 (24.000, 27.090) |
| XGBoost (time series) | 2 |  | 0 | 0.037 (0.035, 0.039) | 0.999 (0.999, 0.999) | 26.100 (24.641, 27.818) |
| XGBoost (time series) | 4 |  | 0 | 0.037 (0.035, 0.039) | 0.999 (0.999, 0.999) | 25.882 (24.381, 27.653) |
| XGBoost (time series) | 6 |  | 0 | 0.038 (0.036, 0.040) | 0.999 (0.999, 0.999) | 25.455 (24.000, 27.090) |
| XGBoost (time series) | 8 |  | 0 | 0.038 (0.036, 0.040) | 0.999 (0.999, 0.999) | 25.316 (23.938, 26.933) |
| XGBoost (time series) | 10 |  | 0 | 0.038 (0.036, 0.040) | 0.999 (0.999, 0.999) | 25.247 (23.691, 26.778) |
| XGBoost (time series) | 12 |  | 0 | 0.038 (0.035, 0.040) | 0.999 (0.999, 0.999) | 25.455 (24.063, 27.249) |
| XGBoost (time series) | 12 |  | 1 | 0.039 (0.037, 0.042) | 0.999 (0.999, 0.999) | 24.510 (23.038, 25.954) |
| XGBoost (time series) | 14 |  | 0 | 0.038 (0.036, 0.040) | 0.999 (0.999, 0.999) | 25.455 (23.814, 27.169) |
| XGBoost (time series) | 16 |  | 0 | 0.038 (0.036, 0.041) | 0.999 (0.999, 0.999) | 25.042 (23.570, 26.624) |
| XGBoost (time series) | 18 |  | 0 | 0.038 (0.036, 0.040) | 0.999 (0.999, 0.999) | 25.247 (23.691, 26.855) |
| XGBoost (time series) | 20 |  | 0 | 0.038 (0.036, 0.040) | 0.999 (0.999, 0.999) | 25.525 (24.126, 27.090) |
| XGBoost (time series) | 22 |  | 0 | 0.038 (0.036, 0.040) | 0.999 (0.999, 0.999) | 25.316 (23.876, 26.933) |
| XGBoost (time series) | 24 |  | 0 | 0.038 (0.036, 0.040) | 0.999 (0.999, 0.999) | 25.385 (23.938, 27.011) |
| XGBoost (time series) | 24 |  | 1 | 0.039 (0.037, 0.041) | 0.999 (0.999, 0.999) | 24.641 (23.155, 26.322) |
| XGBoost (time series) | 26 |  | 0 | 0.038 (0.035, 0.040) | 0.999 (0.999, 0.999) | 25.596 (24.063, 27.329) |
| XGBoost (time series) | 28 |  | 0 | 0.038 (0.035, 0.040) | 0.999 (0.999, 0.999) | 25.596 (24.000, 27.249) |
| XGBoost (time series) | 30 |  | 0 | 0.037 (0.035, 0.039) | 0.999 (0.999, 0.999) | 25.810 (24.445, 27.409) |
| XGBoost (time series) | 32 |  | 0 | 0.037 (0.035, 0.039) | 0.999 (0.999, 0.999) | 25.810 (24.381, 27.571) |
| XGBoost (time series) | 34 |  | 0 | 0.037 (0.035, 0.039) | 0.999 (0.999, 0.999) | 26.100 (24.575, 27.818) |
| XGBoost (time series) | 36 |  | 0 | 0.038 (0.035, 0.040) | 0.999 (0.999, 0.999) | 25.667 (24.126, 27.249) |
| XGBoost (time series) | 38 |  | 0 | 0.037 (0.035, 0.039) | 0.999 (0.999, 0.999) | 25.954 (24.510, 27.571) |
| XGBoost (time series) | 40 |  | 0 | 0.037 (0.035, 0.039) | 0.999 (0.999, 0.999) | 25.882 (24.381, 27.571) |
| XGBoost (time series) | 42 |  | 0 | 0.037 (0.034, 0.039) | 0.999 (0.999, 0.999) | 26.322 (24.840, 28.326) |
| XGBoost (time series) | 44 |  | 0 | 0.037 (0.035, 0.040) | 0.999 (0.999, 0.999) | 25.738 (24.316, 27.490) |
| XGBoost (time series) | 46 |  | 0 | 0.038 (0.036, 0.040) | 0.999 (0.999, 0.999) | 25.667 (24.189, 27.169) |
| XGBoost (time series) | 48 |  | 0 | 0.037 (0.035, 0.039) | 0.999 (0.999, 0.999) | 26.100 (24.575, 27.902) |
| XGBoost (time series) | 48 |  | 1 | 0.038 (0.036, 0.040) | 0.999 (0.999, 0.999) | 25.525 (24.000, 27.169) |
| iTransformer | 12 | Linear | 0 | 0.036 (0.034, 0.038) | 0.999 (0.999, 0.999) | 26.933 (25.385, 28.851) |
| iTransformer | 12 | Linear | 1 | 0.037 (0.034, 0.039) | 0.999 (0.999, 0.999) | 26.322 (24.840, 28.070) |
| iTransformer | 24 | Linear | 0 | 0.035 (0.033, 0.037) | 0.999 (0.998, 0.999) | 27.818 (26.174, 29.675) |
| iTransformer | 24 | Linear | 1 | 0.036 (0.034, 0.039) | 0.999 (0.999, 0.999) | 26.397 (24.907, 28.155) |
| iTransformer | 48 | Linear | 0 | 0.037 (0.035, 0.040) | 0.999 (0.999, 0.999) | 25.882 (24.316, 27.409) |
| iTransformer | 48 | Linear | 1 | 0.035 (0.033, 0.038) | 0.999 (0.999, 0.999) | 27.249 (25.525, 29.211) |

### A.5 Sensitivity Analysis: different imputation strategies for DNN models

For shorter time series (patients have less than 48 hours of history), we performed first observation carried backward imputation for the remaining missing values after linear interpolation, compared with median imputation as a sensitivity analysis. We used vanilla transformer with 12, 24 and 48 hours of lookback. No significant difference was observed across different configurations.

**Figure S2. Area under the Receiver Operating Characteristic Curve (AUROC) (Panel A), Sensitivity (Panel B), and False Alarm Ratio (Panel C) of Transformer model with different interpolation methods. The model used data from 12, 24, 48 hours of lookback period. The error bar indicates the 95% confidence interval.**

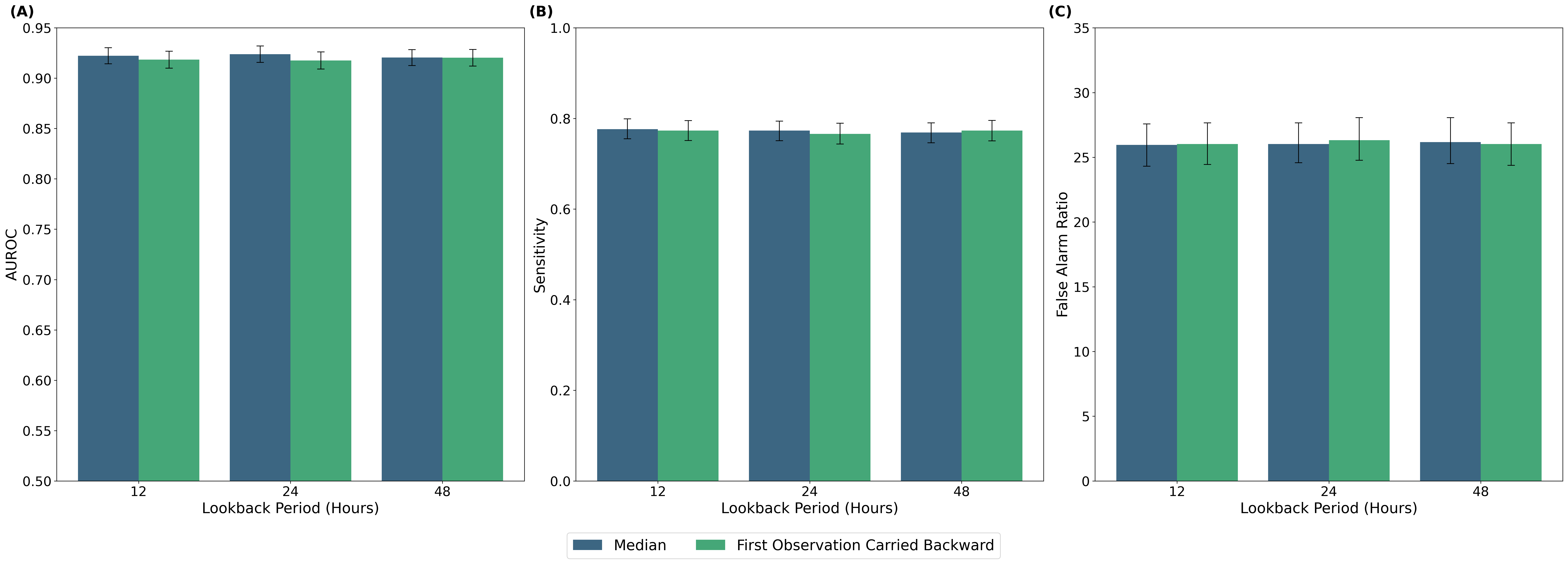
